# Acute Lung injury evolution in Covid-19

**DOI:** 10.1101/2020.08.09.20170910

**Authors:** Doglioni Claudio, Ravaglia Claudia, Rossi Giulio, Dubini Alessandra, Pedica Federica, Piciucchi Sara, Vizzuso Antonio, Pecciarini Lorenza, Stella Franco, Maitan Stefano, Agnoletti Vanni, Gamberini Emiliano, Russo Emanuele, Puglisi Silvia, Arcadu Antonella, Donati Luca, Di Cesare Simona, Grosso Carmela, Poletti Giovanni, Sambri Vittorio, Fabbri D Elisabetta, Pizzolo Giovanni, Ugel Stefano, Bronte Vincenzo, Wells U Athol, Chilosi Marco, Poletti Venerino

**Author notes:** Dual first authorship (Doglioni Claudio). **Corresponding author information:** Claudia Ravaglia, MD. Member of European Reference Network LUNG, ^2^Pulmonology Unit, Thoracic Diseases Department. G.B. Morgagni Hospital, Forlì, Italy.

## Abstract

**Background:** Pathogenesis of Coronavirus disease 2019 (Covid-19) is poorly understood. Most histologic studies come from post-mortem analysis, with existing data indicating that histologic features of acute respiratory distress syndrome are typically present in fatal cases. However, this observation may be misleading, due to confounding factors in pre-terminal disease, including injury resulting from prolonged mechanical ventilation. Ante-mortem lung biopsy may provide major pathogenetic insights, potentially providing a basis for novel treatment approaches.

**Aim:** This comparative, multicenter, prospective, observational study was planned to identify ante-mortem histological profile and immunohistochemical features of lung tissue in patients with Covid-19 in early and late phases of the disease, including markers of inflammatory cells and major pathways involved in the cytokine storm triggering.

**Methods:** Enrolled patients underwent lung biopsy, according to the study protocol approved by local Ethical Committee, either within 15 days of the first symptoms appearing (early phase) or after >15 days (more advanced disease). Key exclusion criteria were excessive or uncorrectable bleeding risk and cardiovascular disease with heart failure. Lung samples were obtained by conventional transbronchial biopsy, trans-bronchial lung cryobiopsy or surgical lung biopsy.

**Results:** 23 patients were enrolled: 12 patients underwent lung biopsy within 15 days and 11 patients more than 15 days after the onset of symptoms. Early biopsies were characterized by spots of patchy acute lung injury (ALI) with alveolar type II cells hyperplasia and significant vascular abnormalities (disordered angiogenesis with alveolar capillary hyperplasia, luminal enlargement and thickened walls of pulmonary venules, perivascular CD4-T-cell infiltration), with no hyaline membranes. In the later stages, the alveolar architecture appeared disrupted, with areas of organizing ALI, venular congestion and capillary thromboembolic microangiopathy. Striking phenotypic features were demonstrated in hyperplastic pneumocytes and endothelial cells, including the expression of phospho-STAT3 and molecules involved in immunoinhibitory signals (PD-L1 and IDO-1). Alveolar macrophages exhibited macrophage-related markers (CD68, CD11c, CD14) together with unusual markers, such as DC-Lamp/CD208, CD206, CD123/IL3AR.

**Conclusion:** A morphologically distinct “Covid pattern” was identified in the earlier stages of the disease, with prominent epithelial and endothelial cell abnormalities, that may be potentially reversible, differing strikingly from findings in classical diffuse alveolar damage. These observations may have major therapeutic implications, justifying studies of early interventions aimed at mitigating inflammatory organ injury.

## INTRODUCTION

Coronavirus disease 2019 (Covid-19) is an infectious disease caused by a novel coronavirus, SARS-Cov2. Covid-19 infection is characterized by clinical variability, with some patients developing mild symptoms but others rapidly progressing to acute respiratory failure requiring intensive care unit (ICU) treatment^1^. Despite intense investigation, the pathogenesis of Covid-19 is poorly understood and the relevance of pathological information has been recently highlighted. However, many studies are centered on post-mortem analysis^2-5^, with only a handful of reports of ante-mortem biopsy findings^6-8^. Post-mortem analysis has generally disclosed diffuse alveolar damage, as typically observed in the acute respiratory distress syndrome (ARDS). A particular emphasis has been given to the presence of thrombo-embolic disease, suggesting a potential role of endothelial related pathways in the pathogenesis; the presence of microangiopathic lesions, perivascular T-cell infiltration, and endothelial injury is in line with this assumption^2,9,10^. The specificity of these findings is questionable, as similar patterns of thrombo-embolic disease, excessive pro-coagulant activity, and vascular changes are common in ARDS regardless of its etiology^11^. In addition, it is possible that prolonged invasive mechanical ventilation significantly impairs the recognition of early and more specific changes in post-mortem samples, due to a variety of confounding factors (medications, ventilator-associated pneumonia, oxygen damage, bacterial super-infections, autolytic changes, etc.). Thus, ante-mortem lung biopsy in patients with Covid-19 has the potential to pinpoint the initial and late phases of the disease without the influence of confounding factors. Here, we examine the morphologic and immuno-molecular features of a series of patients affected by Covid-19 who underwent lung biopsy, either at an earlier or later stage of the illness. The study was planned because Covid-19 associated features appear to amount to a new and unknown entity and, in the lack of solid data, identification of pulmonary histologic lesions might help to drive treatment decisions or drug selection. Furthermore, biopsies were considered clinically useful in order to exclude or confirm super-infections or non-infectious complications in mechanically ventilated patients. This study was in accordance with regulations issued by the

Helsinki Declaration; the protocol was approved by the Institutional Review Board (IRB) of the Area Vasta Romagna Ethical Committee (prot. 2672) and patients and relatives have provided their informed consent, accepting potential risks and advantages of the proposed procedure.

## METHODS

### Study design

Covid-Histology-2020 was a comparative, multicenter, prospective, investigator-initiated, observational study, conducted from April/06^th^/2020, through April/30^th^/2020, that examined lung biopsies obtained from patients with the diagnosis of Covid-19 across two Centers. A total number of 23 patients were recruited. All clinical and laboratory data are described in Table 1. Twelve patients underwent lung biopsy within 15 days of the clinical onset of symptoms (early-phase, group 1), 11 underwent lung biopsy more than 15 days after the onset of symptoms (late-phase, group 2). Mean time between onset of symptoms and lung biopsy was 10 days and 30 days in the two groups, respectively. Lung samples were obtained by conventional trans-bronchial biopsy in 13 cases, by trans-bronchial lung cryobiopsy in 8^12^, by surgical lung biopsy in 2. Bronchoalveolar lavage (BAL) was performed in 11 patients at early-phase and 8 patients at late-phase. Mean duration of hospitalization was 29 days. For study purposes, vital status and post-operative complications were ascertained at study completion. Bronchoscopy and invasive investigations were done according to safety rules applied in our Hospitals, using personal protection equipment including face shield, gown, gloves, and N-95 respirators.

**Table 1.**
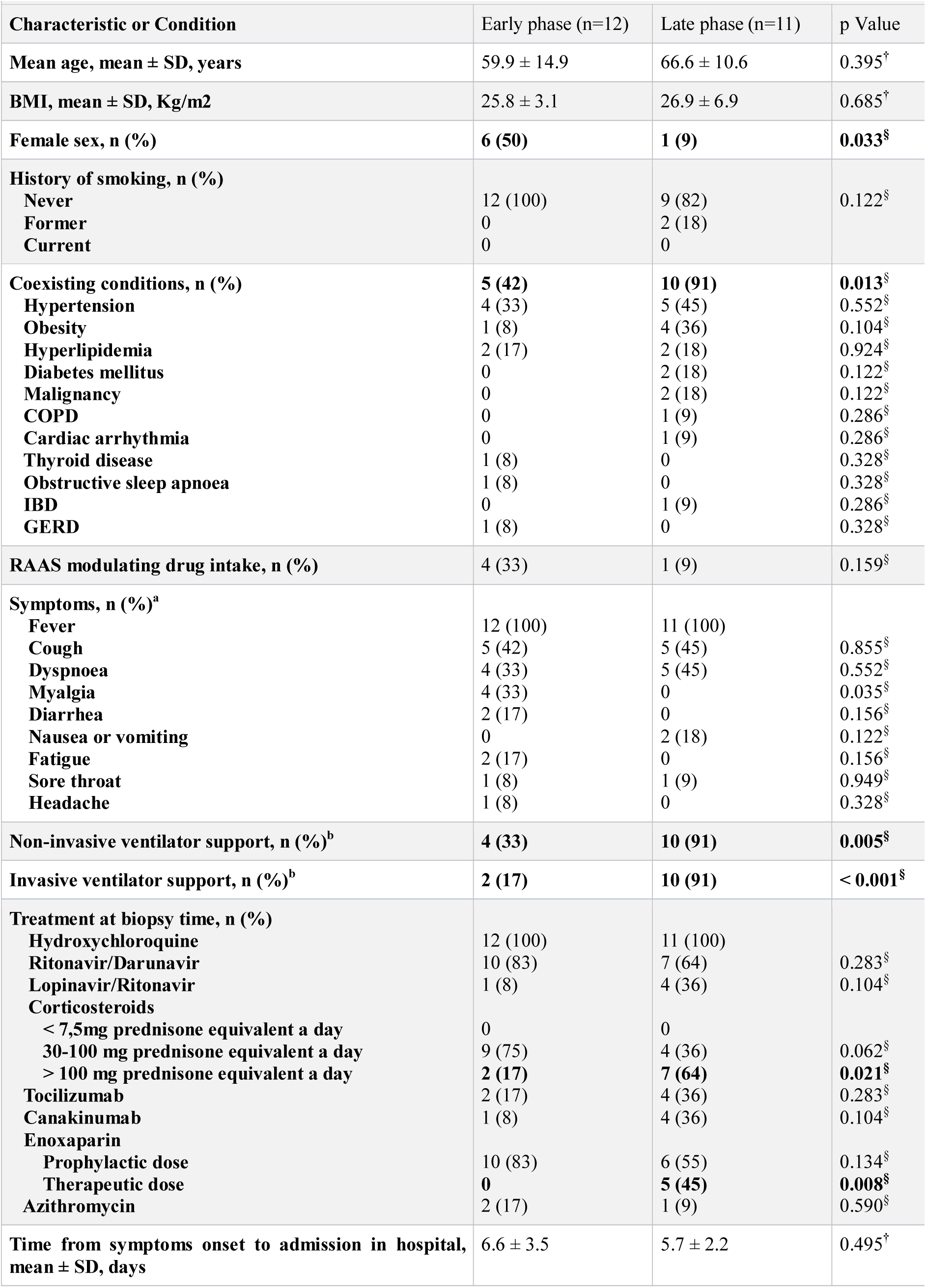
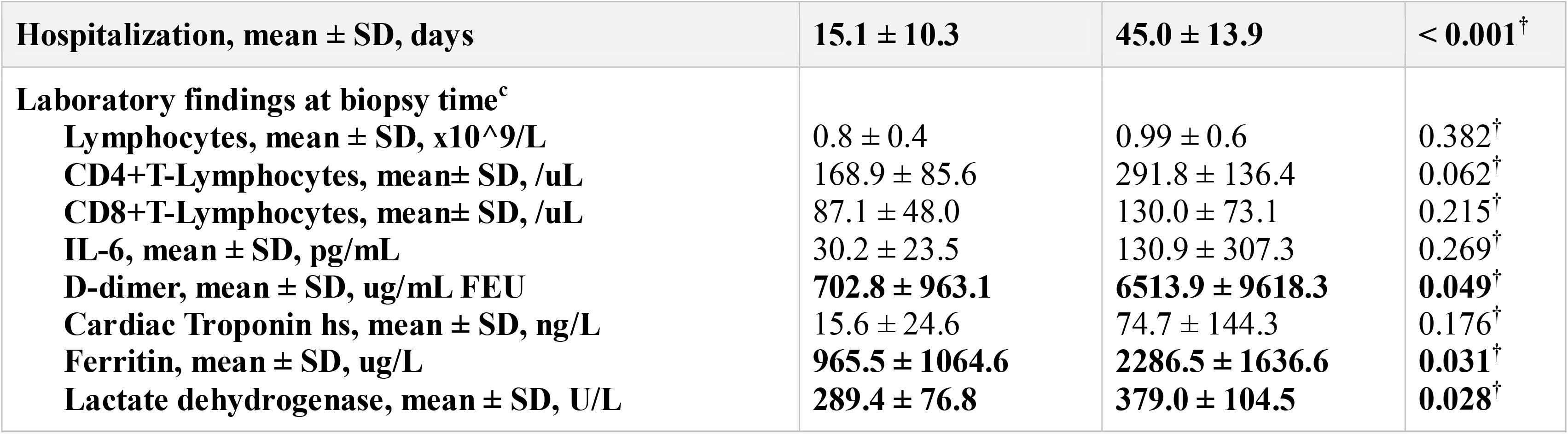
Clinical characteristics and laboratory findings of patients at the time of biopsy.

### Outcomes

Primary aim of the study was the evaluation of the histological profile and the immunohistochemical and molecular features of lung tissues in patients with Covid-19 in the early and late phases of the disease.

### Histopathology and immunohistochemistry

All specimens were fixed in 10% buffered formalin for 12-24 hours, and routinely paraffin-embedded, in order to optimize morphological and immunohistochemical studies on safe non-infective material. Because of the national lockdown, the specimens were reviewed and discussed at virtual meetings on whole slide images (x20-magnification - Aperio/AT2-scanner, Leica). Immunohistochemical markers were applied in order to better recognize and characterize different cell types and phenotypes within the pulmonary microenvironment. The presence of SARS-cov-2 viral genome and IL-6 were investigated using a fluorescence in situ hybridization (FISH) technique and RNA scope technology respectively.

## RESULTS

Clinical characteristics, radiological and laboratory findings are described in Table 1 and supplementary material (eTable 1 in the Supplement).

### Histological patterns

<u>Early phase</u> (12 samples ≤15 days: 7 cryobiopsies, and 5 transbronchial biopsies)

The observed pattern was not the typical diffuse alveolar damage (DAD), observed in ARDS, since hyaline membranes were absent and alveolar epithelial type II cells (AECII) hyperplasia showed a peculiar “patchy” distribution, ranging from cases with isolated small clusters of AECII to wide proliferation of micro-nodular and/or pseudo-papillary sprouts, interposed to variable proportions of normally looking type-I pneumocytes (Table 2). These findings were more evident using CK7 immunostaining (Figure 1). Alveolar epithelial changes were associated with congestion of interstitial capillaries leading to colander-like arrangement, dilatation and tortuosity of postcapillary venules showing thickened, edematous walls, without overt vasculitis nor endotheliitis (Figure 2). On this basis, we categorized this histology picture as *“COVIDpattern”*, distinguishing two scores on the basis of a quantitative evaluation of AECII hyperplasia (Figure 1). Peri-vascular lymphocyte infiltrates were commonly observed. Two cases in early-phase showed thromboembolic microangiopathy involving interstitial capillaries. In early-phase cases, irregular clusters of mononuclear cells were demonstrated within alveolar spaces, characterized by small size and heterogeneous nuclear shape.

**Table 2.**
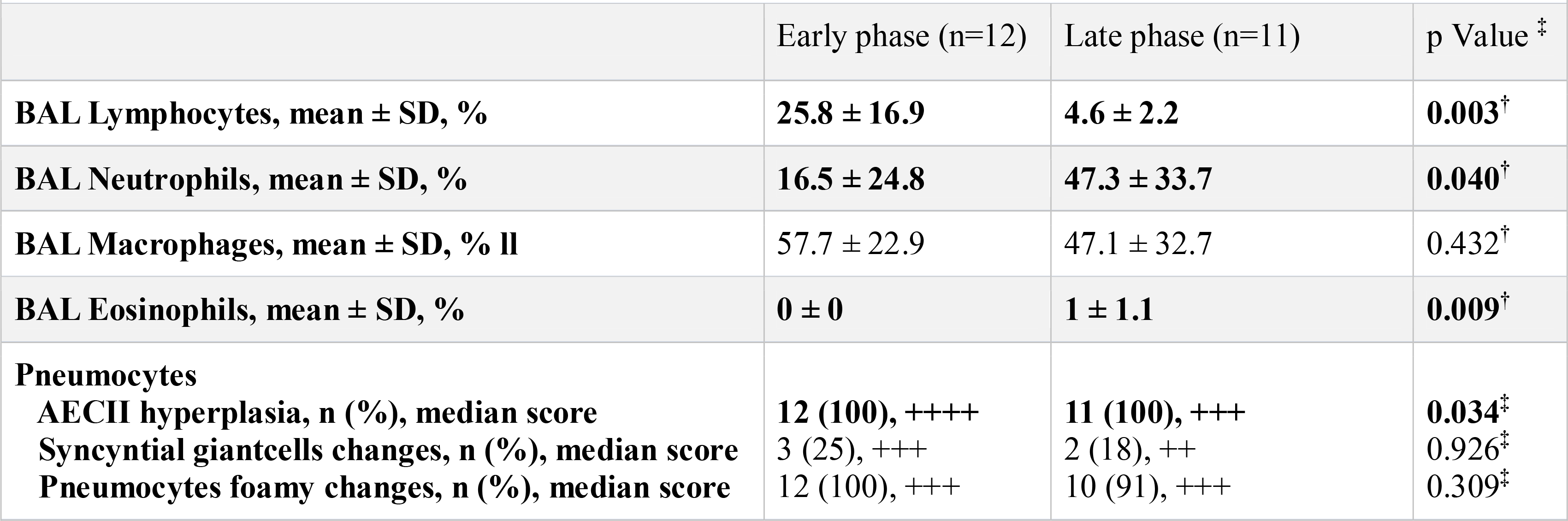
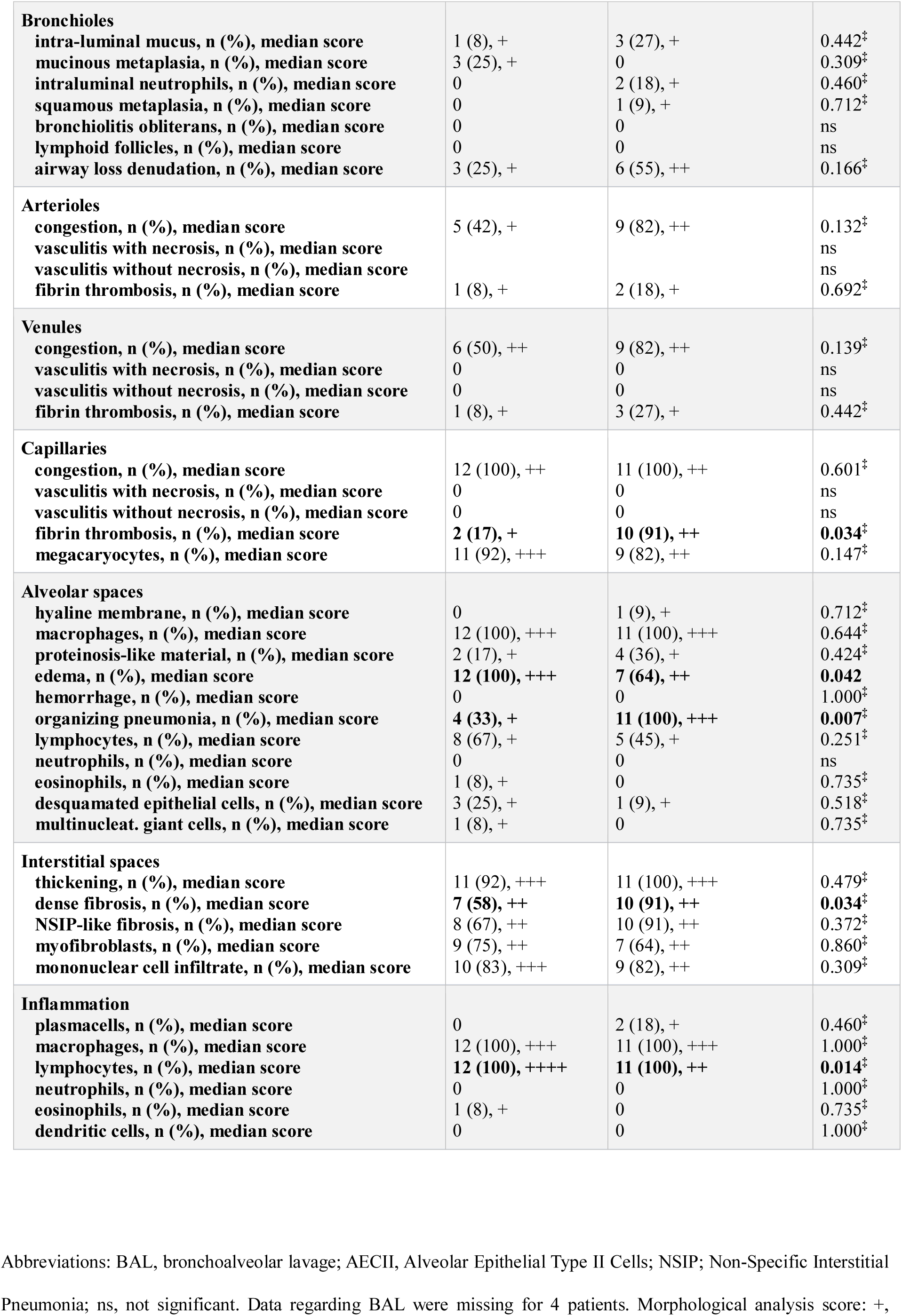
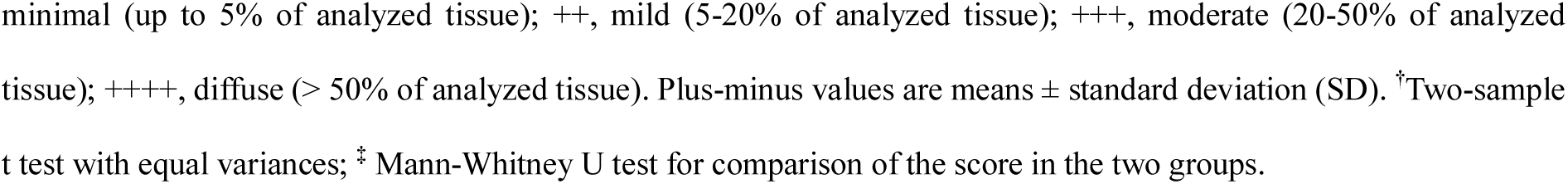
Bronchoscopy data and morphological features through difference compartments.

**Figure 1.**
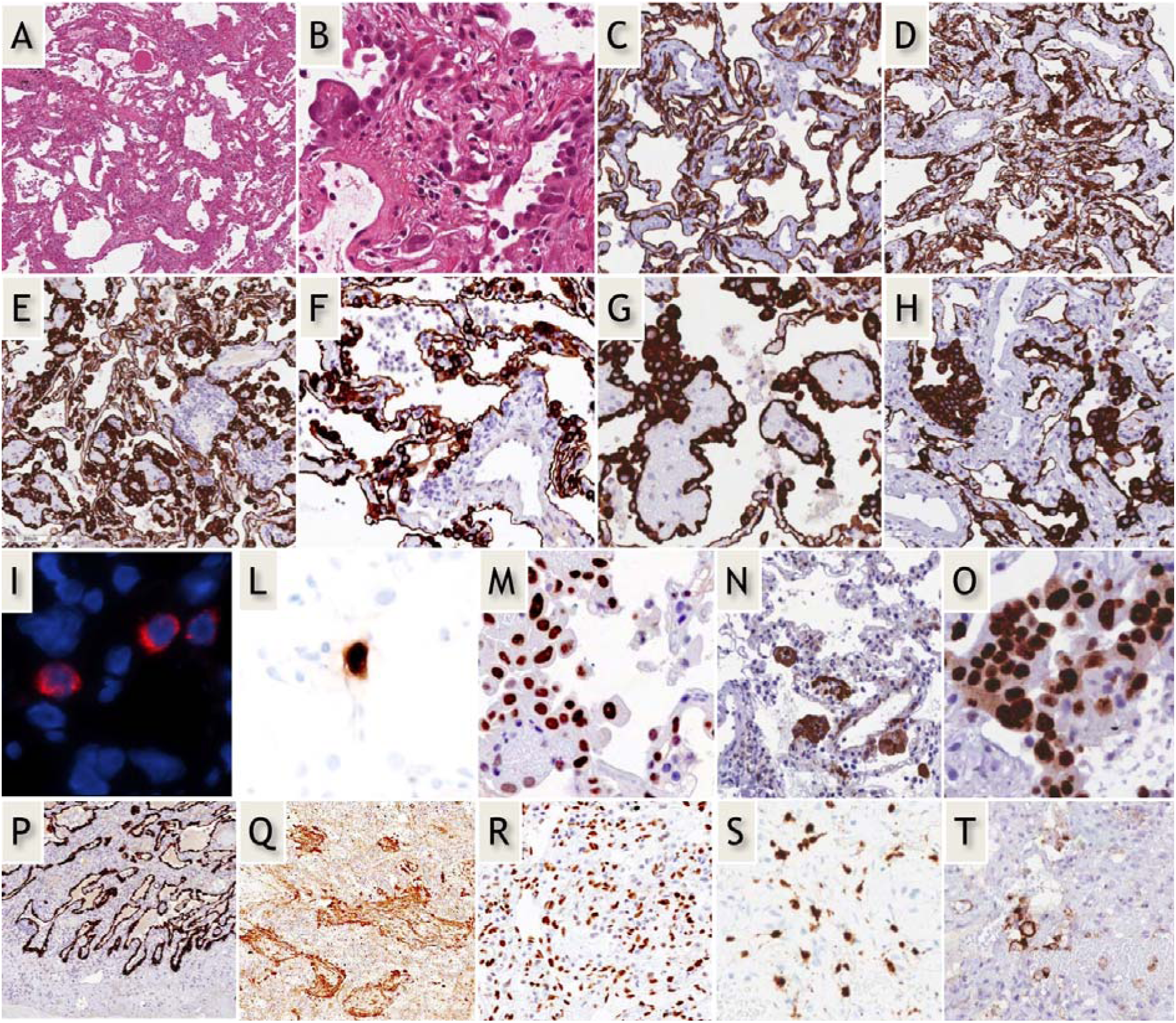
<u>Early-phase COVID-19 pneumonia.</u> H&E (A,B): Parenchymal structure is variably altered by AECII hyperplasia, vascular enlargement and interstitial thickening. CK7 (G-H): AECII form variable small nodules, aggregates and pseudo-papillary sprouts. Grade-1 (C,D) and -2 (E-H) Covid-19 histological patterns were defined by the extent of AEC II hyperplasia. In situ demonstration of AECII infected by SARS-CoV-2 (I): cytoplasmic (red) signals are evidenced in scattered cells recognized as AECII by morphology and location. In situ analysis of IL-6 mRNA expression (L): strong signal is evidenced in scattered AECII. Ph-STAT3 immunohistochemistry (M): strong signal demonstrated in most AECII. TBB3 immunohistochemistry (N): strong signal in AECII. Interstitial dilated spaces are negative. Ki67 immunohistochemistry (O): elevated (>50%) proliferation in AECII. <u>Late-phase Covid-19 pneumonia.</u> CK7 (P): typical DAD-presentation with homogeneous “lepidic” alveolar covering by AECII. TBB3 (Q) strong reaction in myofibroblast-rich areas. phSTAT3 (R): diffuse nuclear expression in AECII, macrophages and stromal cells. IL-6 mRNA in-situ (S): increased numbers of positive cells. PD-L1 (T): negative results in most blood vessels.

**Figure 2.**
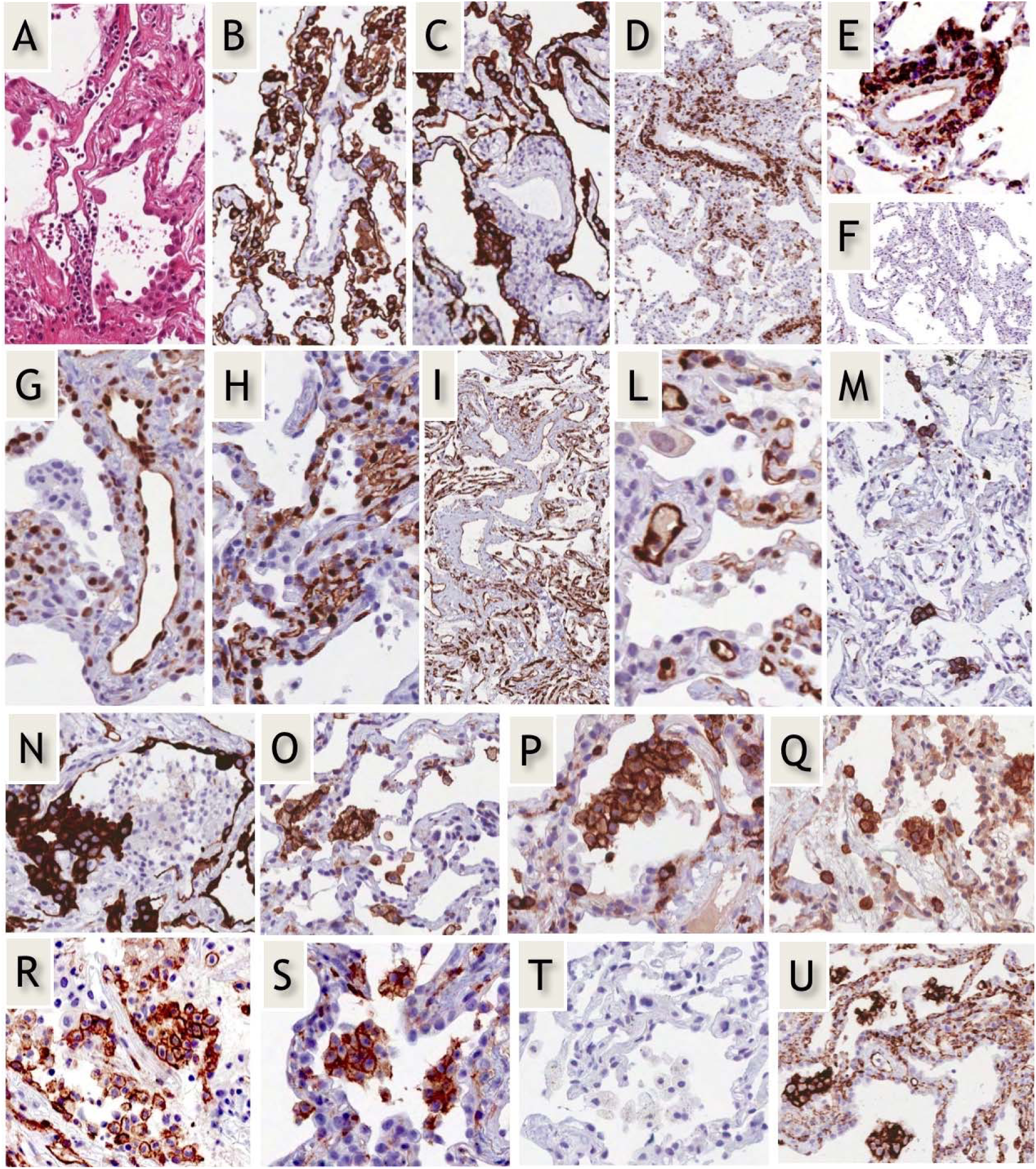
<u>Abnormal morphology and phenotype in enlarged vascular endothelial cells in early-phase Covid-19 pneumonia.</u> H&E (a); CK7 (B,C). lymphocyte infiltration of vascular walls. CD3 (D), CD4 (E), CD8 (F): perivascular lymphocytes mostly exhibit a CD3+, CD4+, CD8-negative immunophenotype. Ph-STAT3 (G): strong nuclear expression in endothelial cells. PD-L1 (H,I), and IDO-1 (L): strong expression in capillaries and venules. CD61 (M): occasional positive megakaryocytes within interstitial capillaries. <u>Immunohistochemical profile of aggregates of alveolar mononuclear cells in early-phase Covid-19 pneumonia.</u> CK7 negative (N), CD11c+ (O), CD4+ (P), CD14+ (Q), CD123+ (R), CD206+ (S), CD303-negative (T), PD-L1+ (U).

<u>Late phase</u> (11 cases >15 days: 1 cryobiopsy, 8 transbronchial biopsies, and 2 open lung biopsies)

In all cases, the alveolar architecture was effaced, with wide areas of parenchyma involved by organizing DAD with diffuse interstitial myofibroblast proliferation (Figure 1). In 7 out of 11 cases, remnants of CK7^+^ epithelial cells were scattered within an hypercellular tissue consisting of interstitial myofibroblasts, inflammatory cells and blood vessels. Hyaline membranes were present in 1 surgical biopsy. Vascular congestion involving arterioles and venules, and thromboembolic microangiopathy of capillaries were observed in 9 out of 11 (82%) of cases in late Covid-19. High levels of D-dimer were found to be associated with presence of capillary fibrin thrombosis (eFigure1 in the Supplement). Residual areas of grade 1 Covid pattern could be observed in 5/11 cases. One surgical biopsy case showed cells infected by cytomegalovirus.

### In situ analysis and immunohistochemistry

Extended immunohistochemical investigations were performed on cryobiopsies, where serial sections were available (15/21- 8 transbronchial cryobiopsy samples; 7 regular transbronchial biopsy samples) and samples obtained by surgical lung biopsies.

#### Early phase

##### a. Epithelial cells

*In situ* analysis using SARS-CoV-2 specific probes evidenced positive cells recognized as AECII by morphology and distribution (Figure 1). The number of FISH^+^ infected cells was higher in score-1 early cases, decreasing where AECII were more prominent. Similar figures were obtained at in situ analysis using a IL-6 specific probe (Figure 1L). AECII exhibited elevated proliferation index (Ki67 >50%; Figure 1O), without morphological evidence of apoptotic bodies, as also confirmed by lack of expression of cleaved caspase-3. Strong nuclear expression of phosphorylated (p)STAT3 was evidenced in all investigated cases (Figure 1M). In control cases, pSTAT3 immunoreactivity was low, confined to scattered AECII and rare blood vessel. AECII strongly expressed Tubulin-beta-3 (TBB3), but interstitial spaces were negative, revealing the rarity/absence of myofibroblasts, at variance with typical DAD (Figure 1N)^13^.

##### b. Macrophages

Irregular clusters of mononuclear cells were demonstrated within alveolar spaces, characterized by small size and heterogeneous nuclear shape. Morphologically, these cells were smaller than typical alveolar macrophages, and exhibited macrophage-related markers (CD68, CD11c, CD14) together with unusual markers, such as DC-Lamp/CD208, CD206, CD123/IL3AR. Several markers (myeloperoxidase, CD303, CD117) were tested to exclude other cell types characterized by constitutive expression of CD123 (i.e. mast cells and plasmacytoid dendritic cells). Expression of PD-L1 was very strong in these clusters, but pSTAT3 and indoleamine 2,3-dioxygenase-1 (IDO-1) were negative (Figure 2).

##### c. Blood vessels

Perivascular CD3^+^ lymphocytes were common in 7/12 cases (Figure 2). Comparable infiltrates were not observed in late-phase and control cases. The T-cell infiltrate was characterized as CD4-positive but negative for the functional and activation markers T-BET, FOXP3, CD25, and CD30. The TH2-related marker GATA3^14^ was expressed by a minority of perivascular lymphocytes (10-20%).A few interstitial PD1^+^, TCF1^+^ lymphocytes were observed, roughly corresponding, for number and location, to CD8^+^ lymphocytes. In all cases, CD56^+^ NK cells, CD20^+^ B-cells and MUM1^+^ plasma cells were either rare or absent. Endothelial cells covering both venules and interstitial capillaries expressed pSTAT3, PD-L1, and IDO-1 at high level (Figure 2). In control cases, the expression of these markers was faint and/or restricted to scattered endothelial cells. CD61 highlighted the presence of an increased number of platelets bordering the surface of small capillaries already in the early phase. Microthrombi could be detected in few early-phase cases (score 2), and in most advanced late-phase cases. Isolated intra-capillary megakaryocytes were demonstrated by CD61-staining in three cases (Figure 2M)

#### Late-phase

In these cases, patterns variability was high. In the two cases where surgical biopsy was available, consistent phenotypic differences from early-phase were observed. AECII hyperplasia was similar to that observed in DAD (Figure 1P-T), and ongoing fibrosis was evidenced by morphology and TBB3 (Figure 2Q). PD-L1 and IDO-1 decorated only scattered vessels and macrophages, which invariably lacked CD123, but strongly expressed pSTAT3 and IL-6.

## DISCUSSION

Acute pneumonias observed in SARS-CoV-2 infection have been morphologically described, as DAD in different phases. Furthermore, thrombi and microthrombi have been recognized as an important morphological component^2,15^. Nevertheless, abnormal coagulation and thrombosis are common in DAD due to a variety of causes, and data from necropsy samples are insufficient for a precise evaluation of the pathogenic mechanisms underlying Covid-19 pneumonia. Several modifications can be determined by injuries related to mechanical ventilation, super-infections or drugs used in the pre-terminal part of the disease. Morphological information regarding the early phases of the disease are therefore needed. In particular, the nature of the cellular targets of infection and their fate, the cells that trigger the cytokine storm, and the role of inflammatory mechanisms occurring within the alveolar microenvironment are not fully defined.

In this study, we demonstrate that the morphological changes do not match any typical DAD pattern in many “early-phase” cases. Clinical and physiological differences between Covid-19 pneumonia and typical DAD have been reported, and the pathological pattern here described is in line with these assumptions*^15,16^*. In particular, the exudative phase with hyaline membranes and alveolar collapse were absent, and the alveolar damage characterized by AECII hyperplasia was patchy, varying from focal AECII hyperplasia to extensive nodular proliferation. In addition, unusual vascular changes, including increased number, dilation and lymphocyte infiltration, were detected in early-phase cases. These observations, taken together with the prominent vascular abnormalities seen in later disease, support the need to move away from traditional ARDS pathways in formulating Covid-19 pathogenesis. This assumption is reinforced by our findings in earlier disease phase, in which unusual vascular changes seems to prevail.

Another relevant difference at odd with ARDS was the scarcity of myofibroblast accumulation in interstitial spaces of Covid-19 cases, as revealed by the lack of staining for the specific marker TBB3 (Figure 2)^13^. On this basis, we described the unique sum of these features as *“Covid* pattern”, sub-divided in two grades on the basis of AECII proliferation.

The findings of this study provide relevant new information. *In situ* hybridization demonstrated the presence of viral genome in cells with morphology and location consistent with AECII (Figure 1i), in agreement with the prevalent angiotensin converting enzyme-2 (ACE2) expression in AECII*^17^*. The proportion of infected epithelial cells was higher in less affected areas, and decreased in areas with high numbers of actively proliferating (Ki67^+^) AECII arguing that AECII do not die following infection but rather receive proliferative signals, as further supported by the absence of apoptotic markers. In this regard, the occurrence of unusual nodules and sprouts of activated AECII appears distinctive for Covid-19.

Another relevant finding of our study is the elevated expression of nuclear pSTAT3 in both AECII and endothelial cells. Moreover, in early phase, also IL-6 mRNA can be demonstrated in AECII but not in inflammatory cells within the alveolar microenvironment (Figure 1L). Imbalanced activation of the NF-kB/STAT3 pathway is generally ascribed to innate immune response following infection^18^ but our study suggests a different scenario whereby the initial local triggering by parenchymal cells might significantly amplify this mechanism and could be a target for specific drugs^19^. Interestingly, in late-phase pneumonia both pSTAT3 and IL-6 are diffusely expressed by a variety of cells in addition to AECII, including macrophages and stromal cells (Figure 1), in line with the increase of cytokine storm in advanced cases^20^. It remains to be determined whether pSTAT3 increase is a consequence of an autocrine/paracrine production of IL-6 or whether the viral infection is directly affecting the phosphorylation of the transcription factor.

The mechanisms accounting for alveolar functional impairment are not related to alveolar loss consequent to AECI lysis and dysepithelization, as commonly observed in DAD, where necrotic epithelial cells form “hyaline membranes”, a morphological marker never observed in early phase cases of our series. Indeed, type-I pneumocytes were morphologically normal and covered large portions of the alveolar surface. The extensive interstitial fibrosis occurring in DAD was not common in Covid-19 early-phase pneumonia, but dominated late-phase cases, which shared pathology patterns described in post-mortem studies. These observations might be related to the use of high positive end-expiratory pressure (PEEP) in these patients, which was advanced to be detrimental^16^.

The number and size of venules were consistently increased in all early cases, and their walls were often infiltrated by CD3^+^, CD4^+^ T-lymphocytes lacking a clear functional polarization, as evidenced by lack of the transcriptional regulators T-BET, FOXP3 and GATA3 expression^21^. Vascular dysfunction is considered a relevant pathogenic component in the disease severity, likely involved in the cascade of coagulation abnormalities eventually leading to thrombo-embolic complications in severely ill patients*^15^*. In a recent study, distinctive vascular changes were demonstrated at autopsy, including severe endothelial injury, intussusceptive neoangiogenesis, micro-thrombi, and microangiopathy*^2^*.

Accordingly, we demonstrated that vascular abnormalities (disordered angiogenesis with luminal enlargement and dilated vessel wall, perivascular T-cell infiltration) are distinctly present from the beginning of covid-19 pneumonia, and can persist through progressive disease stages. Some of these vascular abnormalities (especially ths lymphocytic perivascular infiltration) are mirrored in BAL fluid; in fact, half of the patients in the early phase exhibited BAL lymphocytosis > 20% (Table 2). The frequent lymphocytopenia characterizing Covid-19 patients might thus be caused by different mechanisms involving different T-cell subsets: redistribution to the lung may be relevant for CD4^+^ cells, whereas cytokine-triggered apoptosis and exhaustion be prevalent for CD8^+^ lymphocytes^22^.

Another impressive finding of our study is the demonstration of diffuse and strong expression of PD-L1 and indoleamine 2,3-dioxygenase-1 (IDO-1) in endothelial cells of both interstitial capillaries and venules throughout the early-phases, but either minimal or absent in control samples. The co-regulatory ligand PD-L1 and the tolerogenic enzyme IDO-1 are part of negative feed-back circuits that restrain immune responses and maintain peripheral tolerance, and are involved in immune-escape of tumors from immune attack^23-25^. The endothelial expression of IDO-1 is normally very low or absent in lung tissue but increased after viral infection, likely in response to IFN-g produced by activated lymphocytes^26^.Interestingly, PD-L1 expression is enhanced by IFN-g-JAK1/JAK2-STAT1/STAT2/STAT3-IRF1 axis^27^ and both PD-L1 and IDO-1 are implicated in defense mechanisms from lung injury^28-30^. These findings suggest that the tolerogenic mechanisms previously ascribed to suppressive signals from innate immune responses^31,32^ might be amplified within the pulmonary microenvironment by endothelial expression of PD-L1 and IDO-1 in Covid-19 patients. Interestingly, IDO-1 is also involved in the regulation of vascular remodeling and relaxation, and has a protective role against pulmonary hypertension^33,34^. It is possible to argue that, when hyper-expressed, IDO-1 may exert a relevant role in inducing the pathologically low pulmonary vascular resistance occurring in early-phase COVID-19 pneumonia^35^. These changes might, at least in part, also explain some CT features observed in patients with Covid-19 pneumonia: proximal and distal pulmonary vessel dilatation and tortuosity, predominantly within or surrounding areas of lung opacities^36^. This pulmonary vascular dilation substained by endothelial IDO-1 expression could, at least in part, explain the significant hypoxia (due to ventilatory/perfusion mismatching and increase of intra-alveolar “dead space”) observed in these patients even in the early phase that can be reversed with pronation^35^.

In early-phase pneumonia, we also unveiled an intra-alveolar cell population expressing macrophage markers (CD68, CD14, CD11c) with unusual morphology and phenotype (i.e. positive for CD123, CD208, CD206, and PD-L1). Interestingly, similar phenotypes can be induced in monocytes by pulmonary epithelial cells, determining pro-inflammatory cytokine production^37^. This peculiar “inflammatory-monocyte” population might exert a role in deranging the cytokine milieu in Covid-19 alveolar microenvironment^20^.

In summary, this study provides unprecedented insights into a missing link between the earliest lung alterations and the systemic immune responses in Covid-19 pneumonia, showing a complex scenario where severe derangement of the cross-talk between innate and adaptive immune mechanisms are triggered by viral infection. Tolerogenic and pro-inflammatory signals are produced in the lung by an array of different cell types, including infected AECII, endothelial cells, and specialized monocytes, with simultaneous production of pro-inflammatory and suppressive signals. The balance between the beneficial and detrimental roles of these factors may be crucial for the disease evolution toward either recovery or more severe phases, when oxygen support and ICU become necessary. The close similarity with cancer-related immune impairment can suggest that therapies reverting these altered regulatory circuits might be beneficial also in SARS-cov-2 pneumonia.

## Data Availability

All the data supporting the findings of this study are available from the corresponding author upon reasonable request

## Acknowledgments

We thank Fondazione Cariverona (ENACT Project) for the financial support; AMMP (Associazione Morgagni Malattie Polmonari) for the continuous support; patients themselves who consented to participate in this trial.

## Contributors

CD, CR, MC and VP conceived of and designed the study; CD, CR and MC wrote the paper; CD, CR, GR, AD, FP, SP, AV, EF, MC and VP contributed to data interpretation; all other authors commented on drafts of the paper and contributed to writing of the final version of the manuscript.

## REFERENCES

1. Zhou F, Yu T, Du R, et al. Clinical course and risk factors for mortality of adult inpatients with COVID-19 in Wuhan, China: a retrospective cohort study. Lancet 2020; 395 (10229): 1054-1062. Erratum in: Lancet. 2020;395(10229):1038.

2. Ackermann M, Verleden SE, Kuehnel M, et al. Pulmonary Vascular Endothelialitis, Thrombosis, and Angiogenesis in Covid-19 [published online ahead of print, 2020 May 21]. N Engl J Med. 2020;10.1056/NEJMoa2015432.

3. Xu Z, Shi L, Wang Y, et al. Pathological findings of COVID-19 associated with acute respiratory distress syndrome. Lancet Respir Med. 2020;8:420–422.

4. Carsana L, Sonzogni A, Nasr A, et al. Pulmonary Post-Mortem Findings in a Series of COVID-19 cases from Northern Italy: a Two-Centre descriptive study. Lancet Infect Dis. 2020; S1473-3099(20)30434–5

5. Tian S, Xiong Y, Liu H, et al. Pathological study of the 2019 novel coronavirus disease (COVID-19) through postmortem core biopsies. Mod Pathol. 2020; 33: 1007–1014.

6. Zeng Z, Xu L, Xie XY, et al. Pulmonary Pathology of Early Phase COVID-19 Pneumonia in a Patient with a Benign Lung Lesion [published online ahead of print, 2020 May 6]. Histopathology. 2020;10.1111/his.14138.

7. Cai Y, Hao Z, Gao Y, et al. Coronavirus Disease 2019 in the Perioperative Period of Lung Resection: A Brief Report From a Single Thoracic Surgery Department in Wuhan, People’s Republic of China. J Thorac Oncol. 2020;15:1065–1072.

8. Pernazza A, Mancini M, Rullo E, et al. Early histologic findings of pulmonary SARS-CoV-2 infection detected in a surgical specimen. Virchows Arch. 2020;1–6

9. Varga Z, Flammer AJ, Steiger P, et al. Electron microscopy of SARS-CoV-2: a challenging task - Authors’ reply. Lancet. 2020;395 (10238):e100.

10. Menter T, Haslbauer JD, Nienhold R, et al. Post-mortem examination of COVID19 patients reveals diffuse alveolar damage with severe capillary congestion and variegated findings of lungs and other organs suggesting vascular dysfunction [published online ahead of print, 2020 May 4]. Histopathology. 2020; 10.1111/his.14134.

11. Chambers RC. Procoagulant signalling mechanisms in lung inflammation and fibrosis: novel opportunities for pharmacological intervention? Br J Pharmacol. 2008;153 Suppl 1(Suppl 1):S367–S378.

12. Ravaglia C, Wells AU, Tomassetti S, et al. Diagnostic yield and risk/benefit analysis of trans-bronchial lung cryobiopsy in diffuse parenchymal lung diseases: a large cohort of 699 patients. BMC Pulm Med. 2019;19:16.

13. Chilosi M, Caliò A, Rossi A, et al. Epithelial to mesenchymal transition-related proteins ZEB1, β-catenin, and β-tubulin-III in idiopathic pulmonary fibrosis. Mod Pathol. 2017;30:26–38.

14. Zheng WP, Flavell RA. Pillars Article: The Transcription Factor GATA-2 Is Necessary and Sufficient for Th2 Cytokine Gene Expression in CD4 T Cells. Cell. 1997. 89:587-596. J Immunol. 2016;196:4426–4435

15. Leisman DE, Deutschman CS, Legrand M. Facing COVID-19 in the ICU: vascular dysfunction, thrombosis, and dysregulated inflammation. Intensive Care Med. 2020;46:1105–1108

16. Gattinoni L, Chiumello D, Caironi P, et al. COVID-19 pneumonia: different respiratory treatments for different phenotypes? Intensive Care Med. 2020;46:1099–1102

17. Zhang H, Penninger JM, Li Y, Zhong N, Slutsky AS. Angiotensin-converting enzyme 2 (ACE2) as a SARS-CoV-2 receptor: molecular mechanisms and potential therapeutic target. Intensive Care Med. 2020; 46:586–590.

18. Blanco-Melo D, Nilsson-Payant BE, Liu WC, et al. Imbalanced Host Response to SARS-CoV-2 Drives Development of COVID-19. Cell. 2020;181:1036–1045.e9.

19. Hirano T, Murakami M. COVID-19: A New Virus, but a Familiar Receptor and Cytokine Release Syndrome. Immunity. 2020;52:731–733.

20. Vabret N, Britton GJ, Gruber C, et al. Immunology of COVID-19: Current State of the Science. Immunity. 2020;52:910–941

21. Gagliani N, Huber S. Basic Aspects of T Helper Cell Differentiation. Methods Mol Biol. 2017;1514:19–30.

22. Diao B, Wang C, Tan Y, et al. Reduction and functional exhaustion of T cells in patients with coronavirus disease 2019 (COVID-19). Front Immunol. 2020;11:827.

23. Tumeh PC, Harview CL, Yearley JH, et al. PD-1 blockade induces responses by inhibiting adaptive immune resistance. Nature. 2014;515:568–571.

24. Mondanelli G, Ugel S, Grohmann U, Bronte V. The immune regulation in cancer by the amino acid metabolizing enzymes ARG and IDO. Curr Opin Pharmacol. 2017;35:30–39.

25. Platten M, von Knebel Doeberitz N, Oezen I, Wick W, Ochs K. Cancer Immunotherapy by Targeting IDO1/TDO and Their Downstream Effectors. Front Immunol. 2015;5:673.

26. Bonavente FM, Soto JA, Pizarro-Ortega MS, et al. Contribution of IDO to human respiratory syncytial virus infection. J Leukoc Biol. 2019;106:933–942

27. Garcia-Diaz A, Shin DS, Moreno BH, et al. Interferon Receptor Signaling Pathways Regulating PD-L1 and PD-L2 Expression. Cell Rep. 2017;19:1189-1201. Erratum in: Cell Rep. 2019;29:3766

28. Monaghan SF, Thakkar RK, Heffernan DS, et al. Mechanisms of indirect acute lung injury: a novel role for the coinhibitory receptor, programmed death-1. Ann Surg. 2012;255:158–164.

29. Liu H, Liu L, Visner GA. Nonviral gene delivery with indoleamine 2,3-dioxygenase targeting pulmonary endothelium protects against ischemia-reperfusion injury. Am J Transplant. 2007;7:2291–2300

30. Lomas-Neira J, Monaghan SF, Huang X, Fallon EA, Chung CS, Ayala A. Novel Role for PD-1: PD-L1 as Mediator of Pulmonary Vascular Endothelial Cell Functions in Pathogenesis of Indirect ARDS in Mice. Front Immunol. 2018;9:3030.

31. Schulte-Schrepping J, Reusch N, Paclik D, et al. Suppressive myeloid cells are a hallmark of 1severe COVID-19. MedRxiv. doi: https://doi.org/10.1101/2020.06.03.20119818. Version posted June 5, 2020.

32. Jeannet R, Daix T, Formento R, Feuillard J, François B. Severe COVID-19 is associated with deep and sustained multifaceted cellular immunosuppression. Intensive Care Med. 2020;1–3.

33. Wang Y, Liu H, McKenzie G, et al. Kynurenine is an endothelium-derived relaxing factor produced during inflammation. Nat Med. 2010;16:279–285

34. Xiao Y, Christou H, Liu L, et al. Endothelial indoleamine 2,3-dioxygenase protects against development of pulmonary hypertension. Am J Respir Crit Care Med 2013; 188: 482–491

35. Gattinoni L, Coppola S, Cressoni M, Busana M, Rossi S, Chiumello D. COVID-19 Does Not Lead to a "Typical" Acute Respiratory Distress Syndrome. Am J Respir Crit Care Med. 2020;201:1299–300

36. Lang M, Som A, Mendoza DP, et al. Hypoxaemia related to COVID-19: vascular and perfusion abnormalities on dual-energy CT. Lancet Infect Dis. 2020; S1473-3099(20)30367–4

37. Gazdhar A, Blank F, Cesson V, et al. Human Bronchial Epithelial Cells Induce CD141/CD123/DC-SIGN/FLT3 Monocytes That Promote Allogeneic Th17 Differentiation. Front Immunol. 2017;8:447

